# Analytical performance of lateral flow immunoassay for SARS-CoV-2 exposure screening on venous and capillary blood samples

**DOI:** 10.1101/2020.05.13.20098426

**Authors:** Margaret A. Black, Guomiao Shen, Xiaojun Feng, Wilfredo F. Garcia Beltran, Yang Feng, Varshini Vasudevaraja, Douglas Allison, Lawrence H. Lin, Tatyana Gindin, Michael Astudillo, Diane Yang, Mandakolathur Murali, A. John Iafrate, George Jour, Paolo Cotzia, Matija Snuderl

## Abstract

**Objectives:** Numerous serologic immunoassays have been launched to detect antibodies to SARS-CoV-2, including rapid tests. Here, we validate use of a lateral flow immunoassay (LFI) intended for rapid screening and qualitative detection of anti-SARS-CoV-2 IgM and IgG in serum, plasma, and whole blood, and compare results with ELISA. We also seek to establish the value of LFI testing on blood obtained from a capillary blood sample.

**Methods:** Samples collected by venous blood draw and capillary finger stick were obtained from patients with SARS-CoV-2 detected by RT-qPCR and control patients negative for SARS-CoV-2. Samples were tested with the 2019-nCoV IgG/IgM Detection Kit (Colloidal Gold) lateral flow immunoassay, and antibody calls were compared with results obtained by ELISA.

**Results:** The Biolidics LFI kit shows clinical sensitivity of 92% at 7 days after PCR diagnosis of SARS-CoV-2 on venous blood. Test specificity was 92% for IgM and 100% for IgG. There was no significant difference in detecting IgM and IgG with Biolidics LFI and ELISA at D0 and D7 (p=1.00), except for detection of IgM at D7 (p=0.04). Finger stick whole blood of SARS-CoV-2 patients showed 93% sensitivity for antibody detection.

**Conclusions:** Clinical performance of Biolidics 2019-nCoV IgG/IgM Detection Kit (Colloidal Gold) is comparable to ELISA and showed consistent results across different sample types. Furthermore, we show that capillary blood obtained by finger stick shows similar sensitivity for detecting anti-SARS-CoV-2 IgM and IgG antibodies as venous blood samples. This provides an opportunity for decentralized rapid testing in the community and may allow point-of-care and longitudinal self-testing for the presence of anti-SARS-CoV-2 antibodies.

## Background

The novel severe acute respiratory coronavirus, SARS-CoV-2, emerged in Wuhan, China in December of 2019 [1]. The disease caused by SARS-CoV-2, COVID-19, rapidly spread across the globe and was declared a pandemic by the World Health Organization (WHO) on March 11, 2020 [2]. COVID-19 has caused significant morbidity and mortality worldwide, with over 4 million confirmed cases and 284,536 deaths attributed to the disease as of May 11, 2020 [3]. In the United States alone, there are over 1 million confirmed cases and more than 80,000 deaths attributed to COVID-19 [4].

Reverse-transcriptase polymerase chain reaction (RT-PCR) techniques performed with nasopharyngeal samples are the mainstay for diagnosing acute infection with SARS-CoV-2, and multiple molecular testing modalities are now available [5]. However, serologic testing to determine recent infections and potential immunity remain limited. Serologic testing for IgM and IgG antibodies is a useful adjunct for clinical decision making [6] and has important implications for public health and policy decisions [7]. Serology is cost-efficient, fast, simple to perform, and does not require additional materials such as nasopharyngeal swabs and viral transport media required for many PCR-based molecular testing platforms, which have been scarce during the pandemic. With no available vaccine and limited treatment options for COVID-19, the development and validation of rapid serologic testing is urgently required. Serologic assessment provides valuable information on past exposure, although the protective effect of anti-SARS-CoV2 antibodies remains unknown.

As demands for laboratory testing have increased exponentially, commercial vendors are developing *in vitro* diagnostics for detection of SARS-CoV2, and many are applying for and obtaining emergency use authorization (EUA) from the Food and Drug Administration (FDA) [7]. In addition to RT-PCR diagnostics, a wide range of serologic immunoassays have been developed, including rapid tests. Early on in this pandemic, the FDA approached serologic testing with limited oversight and did not require EUA, in contrast to requirements for molecular assays. Under the FDA Policy D, providers or manufacturers of serologic tests had to notify the FDA about tests, but those tests were not subject to review. This led to an increasing number of serologic assays listed under policy D, and rapid implementation of tests in the field. However, highly variable sensitivity and specificity of these assays for COVID-19 immunity quickly led to the recognition that regulatory oversight or robust internal validation is required. Tests approved by the FDA under EUA must be verified before they are widely used for clinical diagnosis and decision-making. In the absence of FDA approval, internal rigorous validation of intended assays with establishing proper thresholds is necessary.

Herein, we describe clinical validation of a new lateral flow immunoassay (LFI) test intended for rapid screening and qualitative detection of anti-SARS-CoV-2 IgM and IgG in serum, plasma, and whole blood. We also sought to establish the value of LFI testing on capillary blood obtained from a finger stick sample, since there are currently no FDA EUA approved assays for capillary blood. Plasma, serum, whole blood, and capillary blood (finger stick) samples from patients diagnosed with COVID-19 by PCR were tested with the 2019-nCoV IgG/IgM Detection Kit (Colloidal Gold) (Biolidics Ltd.), and the results were correlated with those obtained by enzyme-linked immunosorbent assay (ELISA), the gold standard for serologic detection of antibodies.

## Materials and Methods

### Study design

This retrospective study assessed the sensitivity and specificity of a commercially available lateral flow immunoassay for detection of IgM and IgG antibodies specific to SARS-CoV-2. Samples from 62 patients used in this study were either discarded clinical samples collected for routine laboratory tests or IRB approved research samples collected at NYU Langone Medical Center (S16-00122).

### Blood samples

Whole blood, plasma, or serum samples collected by venous blood draw and capillary blood collected via finger stick were used. Patients were defined as SARS-CoV-2 positive if RT-qPCR (cobas, Roche FDA EUA) performed on nasopharyngeal samples was positive for SARS-CoV-2 sequence. Plasma and venous whole blood samples were collected from hospitalized SARS-CoV-2 positive patients on the day of the initial positive nasopharyngeal PCR test (D0; n=24) or ≥7 days (D7; n=26) after the positive PCR test (D7). For 11 patients, serum samples at two time points separated by 7 days were available (D0 and D7). Plasma samples from hospitalized patients with SARS-CoV-2 not detected in nasopharyngeal sample by PCR, but positive for other respiratory viruses (human metapneumovirus, rhinovirus/enterovirus, coronavirus NL63, Influenza A H1N1), were also collected at D0 (n=4). In addition, plasma samples of presumably SARS-CoV-2 negative patients hospitalized in January-April of 2019 (pre-SARS-CoV-2 era) that were previously collected and stored at −80°C were used (n=20). Samples collected for validation of capillary whole blood samples were obtained by finger stick from 14 patients who had recovered from COVID-19 at 18-46 days after positive SARS-CoV-2 positive RT-qPCR testing.

### Clinical SARS-CoV-2 testing

Nasopharyngeal samples were tested for clinical care using the cobas® SARS-CoV-2 molecular diagnostic test (Roche) following the manufacturer’s instructions per FDA approved EUA protocol.

### Biolidics Lateral Flow Immunoassay

Venous plasma, serum, whole blood, or capillary (finger stick) blood was used for the 2019-nCoV IgG/IgM Detection Kit (Colloidal Gold) (Biolidics Ltd., Singapore) following the manufacturer’s instructions. This lateral flow immunoassay test is based on solid-phase immunochromatography. Briefly, one drop (20 μL) of room temperature (18-28 °C) plasma, serum, whole blood, or capillary finger stick blood was added to the sample well of a test cassette using the provided dropper, and then 3 drops of diluent were added into the same sample well. Results were read after 10 minutes by assessing visual color changes in the testing strip corresponding to IgM, IgG, and control protein regions, and the image was electronically documented. Each test was reviewed by two observers. The IgM or IgG band was called positive only if identified by both observers independently.

Any intensity of red color change in the IgM and IgG test regions, together or alone, was considered a positive result (Figure 1). Plasma, serum, and whole blood samples were tested for anti-SARS-CoV-2 IgM and IgG antibodies at D0 or D7, and capillary finger stick samples were obtained more than 14 days from both symptom onset (range, 19-61 days; median, 32 days) and PCR diagnosis (range, 18-46 days; median, 30.5 days).

**Figure 1.**
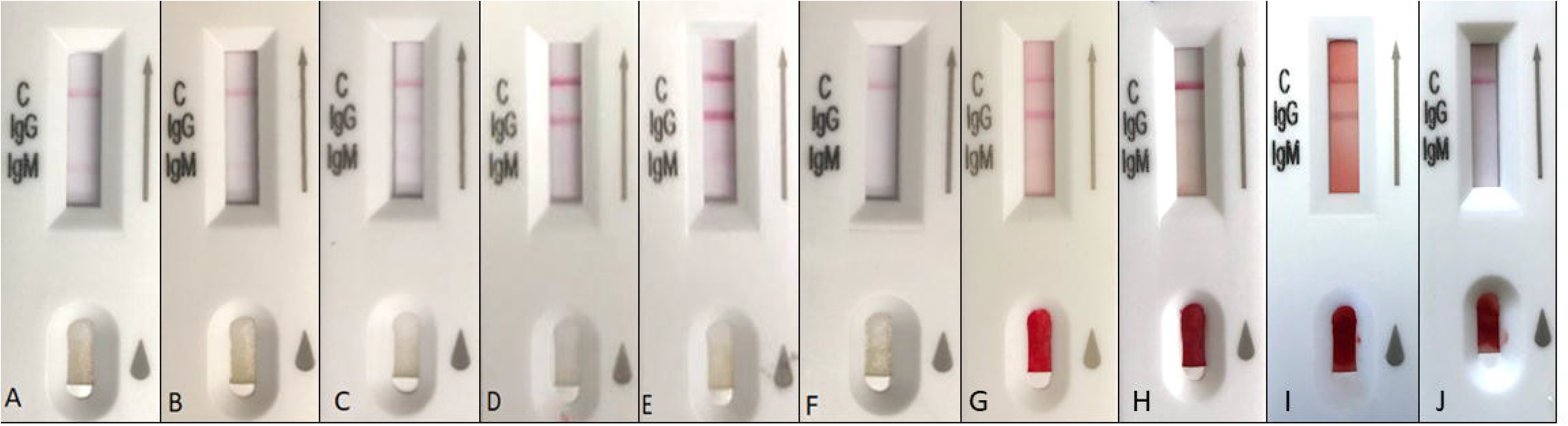
Visual interpretation of 2019-nCoV IgG/IgM Detection Kit (Colloidal Gold). Any intensity of red color change in the IgM and IgG test regions is considered a positive result. Plasma results: A. IgM (+) IgG (-), B. IgM (+) IgG (-), C. IgM (+) IgG (+), D. IgM (-) IgG (+), E. IgM (+) IgG (+), F. IgM (-) IgG (-). Capillary blood results: G. IgM (+) IgG (+), H. IgM (-) IgG (+), I. IgM (-) IgG (+), J. IgM (-) IgG (-).

### Enzyme-linked immunosorbent assay (ELISA)

The MGH/Ragon COVID-19 IgG, IgA, and IgM ELISA, an in-house ELISA developed by Massachusetts General Hospital (Boston, MA) and Ragon Institute of MGH, MIT, and Harvard (Cambridge, MA), was performed on plasma and serum specimens for quantitative assessment of IgG, IgA, and IgM antibodies that target SARS-CoV-2 receptor binding domain (RBD). Sequential specimens of 9 patients were analyzed at two time points 7 days apart, and 6 additional specimens were analyzed at D0 (total D0, n=15; total D7, n=9). In addition, 24 specimens negative for SARS-CoV-2 were assessed by ELISA. Quantitation of antibodies was performed by interpolating O.D. values to a standard curve consisting of an anti-SARS-CoV-1/2 monoclonal antibody (CR3022) in IgG, IgA, and IgM isotypes. RBD and CR3022-IgG were kindly produced and provided by Jared Feldman, Tim M. Caradonna, and Blake M. Hauser from the laboratory of Dr. Aaron Schmidt (Ragon Institute). CR3022 reference sequences, CR3022-IgA and CR3022-IgM isotypes, and ELISA protocols were kindly provided by Stephanie Fischinger, Caroline Atyeo, and Matthew Slein from the laboratory of Dr. Galit Alter (Ragon Institute). ELISA protocols were developed and optimized with the laboratory of Dr. Alejandro Balazs (Ragon Institute). Automation was performed on a QUANTA-Lyser 3000 (Inova).

### Statistical Analysis

To compare Biolidics and ELISA in detecting antibodies at D0 and D7, a pair-wise two-sample t-test was conducted (Table 3). Pair-wise two-sample t-test was also conducted to compare plasma and whole blood samples with Biolidics (Table 4). Exact 95% confidence intervals (Clopper–Pearson Method for Binomial Proportions) of percentage positivity for anti-SARS-CoV-2 IgM and IgG were calculated in patients with SARS-CoV-2 assessed by Biolidics (Table 1B) and in patients with SARS-CoV-2 assessed by ELISA (Table 3C). The sensitivity of IgM and IgG detection with capillary samples in patients with SARS-CoV-2 was also calculated (Table 4D). P-values lower than 0.05 were considered statistically significant. The calculations were performed using R software.

## Results

### Clinical Agreement: Clinical Sensitivity and Specificity

Results of a commercially available serologic LFI for detecting anti-SARS-CoV-2 IgM and IgG antibodies were evaluated using plasma and serum samples of 39 PCR-positive patients with SARS-CoV-2 and negative control samples from 24 patients. Of the 39 PCR-positive patients, 13 patients were tested by LFI on the day of PCR diagnosis (D0), 15 patients were tested at 7 days post PCR test (D7), and 11 patients were sequentially assessed at both D0 and D7 (Table 1A). Four negative control samples were obtained in March 2020 from patients with negative nasopharyngeal PCR for SARS-CoV-2, and 20 negative control samples were archived samples of patients hospitalized in early 2019 before the emersion of SARS-CoV-2 (Table 2A).

To assess the sensitivity of LFI at various time points after initial diagnosis, plasma samples were grouped by number of days since initial detection of SARS-CoV-2 on nasopharyngeal sample. D0 indicates that LFI was performed using plasma from the same day as the initial nasopharyngeal sample, and D7 indicates that LFI was performed using plasma from approximately one week after the initial nasopharyngeal sample was obtained. The sensitivity for detecting IgM, IgG, and IgM or IgG at D0 was 29%, 21%, and 29%, respectively (Table 1B). Sensitivity at D7 increased to 54% for IgM, 88% for IgG, and 92% for IgM or IgG (Table 1B).

**Table 1A:**
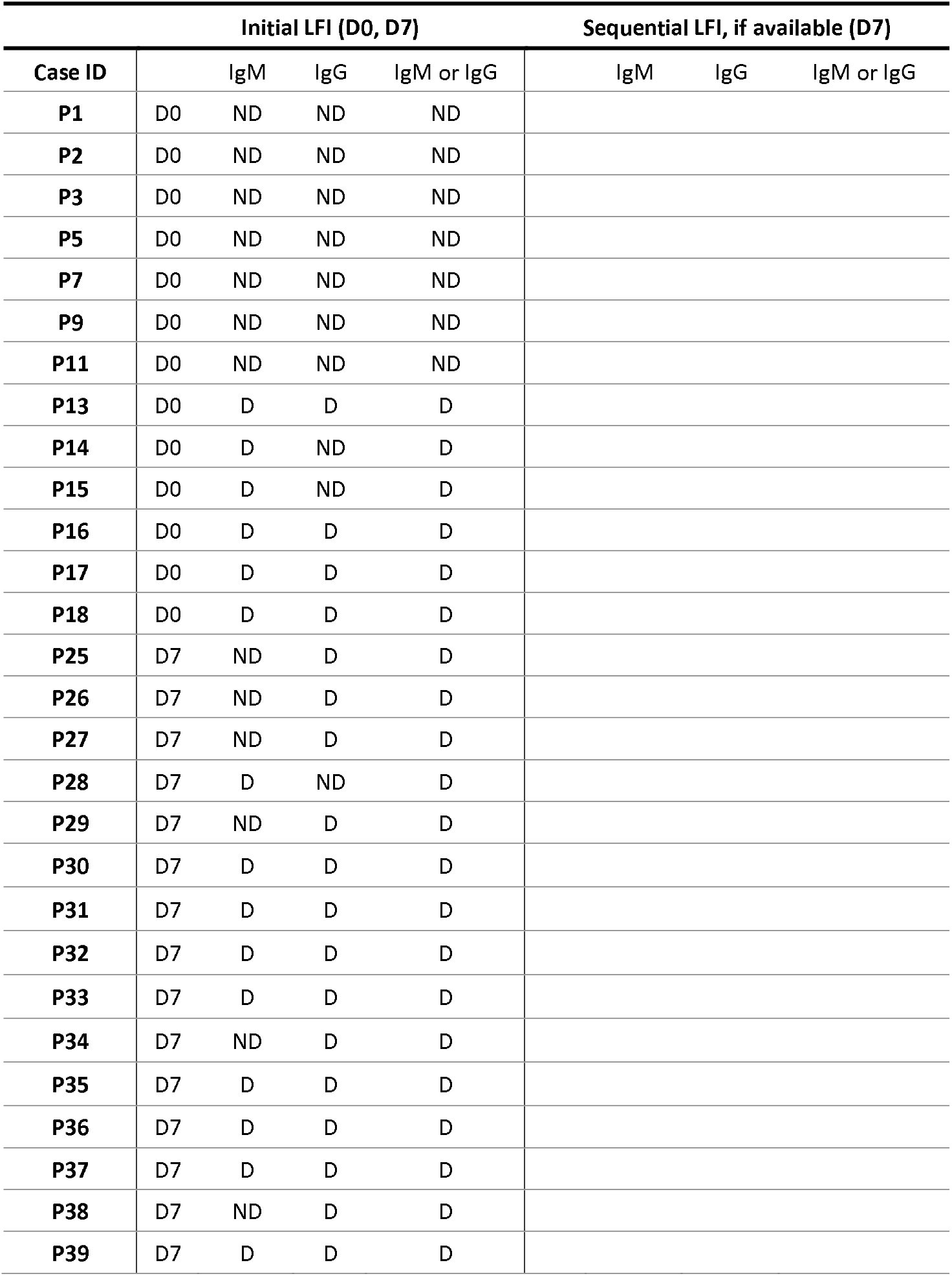
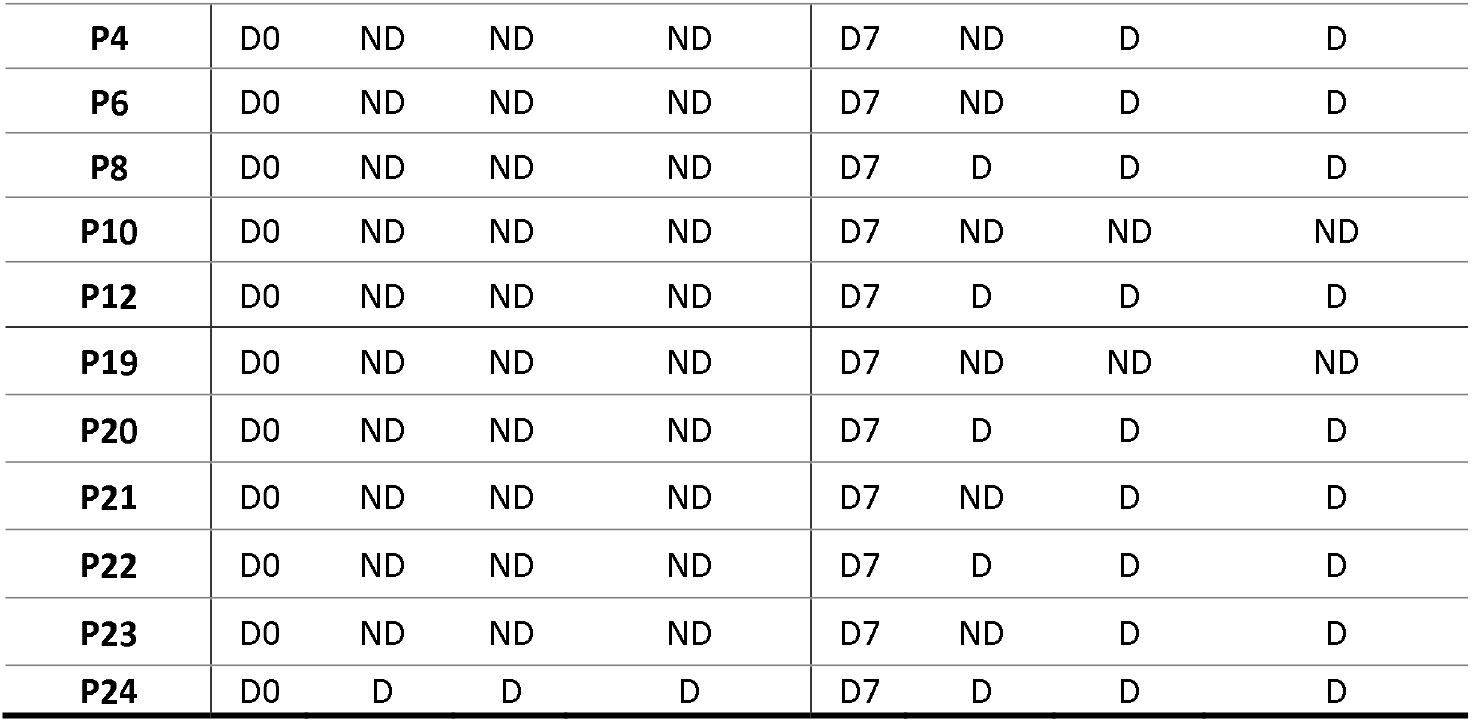
Clinical performance of Biolidics in patients with SARS-CoV-2

| Case ID | Initial LFI (D0, D7) |  |  |  | Sequential LFI, if available (D7) |  |  |
| --- | --- | --- | --- | --- | --- | --- | --- |
|  |  | IgM | IgG | IgM or IgG | IgM | IgG | IgM or IgG |
| P1 | D0 | ND | ND | ND |  |  |  |
| P2 | D0 | ND | ND | ND |  |  |  |
| P3 | D0 | ND | ND | ND |  |  |  |
| P5 | D0 | ND | ND | ND |  |  |  |
| P7 | D0 | ND | ND | ND |  |  |  |
| P9 | D0 | ND | ND | ND |  |  |  |
| P11 | D0 | ND | ND | ND |  |  |  |
| P13 | D0 | D | D | D |  |  |  |
| P14 | D0 | D | ND | D |  |  |  |
| P15 | D0 | D | ND | D |  |  |  |
| P16 | D0 | D | D | D |  |  |  |
| P17 | D0 | D | D | D |  |  |  |
| P18 | D0 | D | D | D |  |  |  |
| P25 | D7 | ND | D | D |  |  |  |
| P26 | D7 | ND | D | D |  |  |  |
| P27 | D7 | ND | D | D |  |  |  |
| P28 | D7 | D | ND | D |  |  |  |
| P29 | D7 | ND | D | D |  |  |  |
| P30 | D7 | D | D | D |  |  |  |
| P31 | D7 | D | D | D |  |  |  |
| P32 | D7 | D | D | D |  |  |  |
| P33 | D7 | D | D | D |  |  |  |
| P34 | D7 | ND | D | D |  |  |  |
| P35 | D7 | D | D | D |  |  |  |
| P36 | D7 | D | D | D |  |  |  |
| P37 | D7 | D | D | D |  |  |  |
| P38 | D7 | ND | D | D |  |  |  |
| P39 | D7 | D | D | D |  |  |  |

|  |  |  |  |  |  |  |  |  |
| --- | --- | --- | --- | --- | --- | --- | --- | --- |
| **P4** | D0 | ND | ND | ND | D7 | ND | D | D |
| **P6** | D0 | ND | ND | ND | D7 | ND | D | D |
| **P8** | D0 | ND | ND | ND | D7 | D | D | D |
| **P10** | D0 | ND | ND | ND | D7 | ND | ND | ND |
| **P12** | D0 | ND | ND | ND | D7 | D | D | D |
| **P19** | D0 | ND | ND | ND | D7 | ND | ND | ND |
| **P20** | D0 | ND | ND | ND | D7 | D | D | D |
| **P21** | D0 | ND | ND | ND | D7 | ND | D | D |
| **P22** | D0 | ND | ND | ND | D7 | D | D | D |
| **P23** | D0 | ND | ND | ND | D7 | ND | D | D |
| **P24** | D0 | D | D | D | D7 | D | D | D |

**Table 1B:** Percentage positivity for anti-SARS-CoV-2 IgM and IgG by Biolidics in patients with SARS-CoV-2

|  |  | **Total,**<br>**n** | **Positive, n** | **Sensitivity**<br>**(%)** | **95% CIs** |
| --- | --- | --- | --- | --- | --- |
| **D0** | IgM+ | 24 | 7 | 29% | 13-51% |
|  | IgG+ | 24 | 5 | 21% | 7-42% |
|  | IgG+ or IgM+ | 24 | 7 | 29% | 13-51% |
| **D7** | IgM+ | 26 | 14 | 54% | 33-73% |
|  | IgG+ | 26 | 23 | 88% | 70-98% |
|  | IgG+ or IgM+ | 26 | 24 | 92% | 75-99% |

\*D, detected; ND, not detected; D0, day 0 after PCR diagnosis; D7, day 7 after PCR diagnosis.

Of 24 negative plasma samples, 22 tested negative for IgM and IgG antibodies (92%) by LFI. Of 4 negative plasma samples obtained in March 2020, no IgM or IgG was detected (Table 2A). Interestingly, of 20 plasma samples obtained from before the SARS-CoV-2 outbreak, 2 samples tested positive for IgM, however no samples tested positive for IgG (Table 2A). The overall specificity of the LFI was 92% for IgM, 100% for IgG, and 92% for IgM and IgG combined (Table 2B).

**Table 2A.** Clinical performance of Biolidics in patients without SARS-CoV-2

| Case ID | SARS-CoV-2<br>PCR | IgM | IgG | IgM or IgG |
| --- | --- | --- | --- | --- |
| N1 | ND | ND | ND | ND |
| N2 | ND | ND | ND | ND |
| N3 | ND | ND | ND | ND |
| N4 | ND | ND | ND | ND |
| N7 | Pre-COVID | D | ND | D |
| N8 | Pre-COVID | ND | ND | ND |
| N9 | Pre-COVID | ND | ND | ND |
| N10 | Pre-COVID | ND | ND | ND |
| N11 | Pre-COVID | ND | ND | ND |
| N12 | Pre-COVID | ND | ND | ND |
| N13 | Pre-COVID | ND | ND | ND |
| N14 | Pre-COVID | ND | ND | ND |
| N15 | Pre-COVID | ND | ND | ND |
| N16 | Pre-COVID | ND | ND | ND |
| N17 | Pre-COVID | ND | ND | ND |
| N18 | Pre-COVID | ND | ND | ND |
| N19 | Pre-COVID | ND | ND | ND |
| N20 | Pre-COVID | ND | ND | ND |
| N21 | Pre-COVID | ND | ND | ND |
| N22 | Pre-COVID | ND | ND | ND |
| N23 | Pre-COVID | ND | ND | ND |
| N24 | Pre-COVID | D | ND | D |
| N25 | Pre-COVID | ND | ND | ND |
| N26 | Pre-COVID | ND | ND | ND |

\* Samples N1-N4 were obtained in patients with respiratory disease in March 2020 who were negative for SARS-CoV-2 by PCR
\* Samples N7-N26 were obtained from patients hospitalized in the pre-COVID19 era (January - April, 2019)
\*D, detected; ND, not detected.

**Table 2B.**
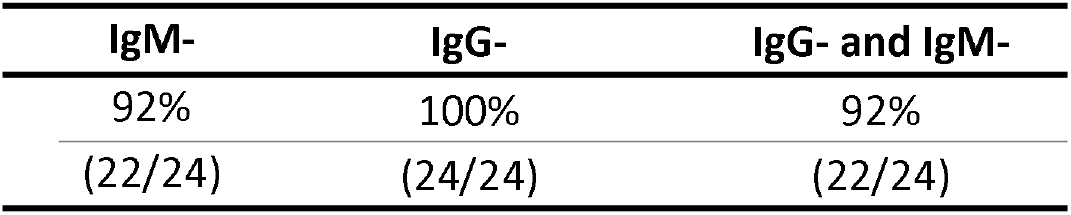
Specificity of IgM and IgG detection by Biolidics in patients with SARS-CoV-2

Eleven patients with SARS-CoV-2 were tested sequentially at D0 and D7. One patient was positive for IgM and IgG antibodies at D0 and also at D7. Of the 10 patients who were negative for antibodies at D0, 4 showed IgM antibodies at D7 and 8 showed IgG antibodies at D7 time point. Overall, 9 of 11 sequentially tested patients showed IgM and/or IgG antibodies at D7 by Biolidics LFI (Figure 2A).

**Figure 2.**
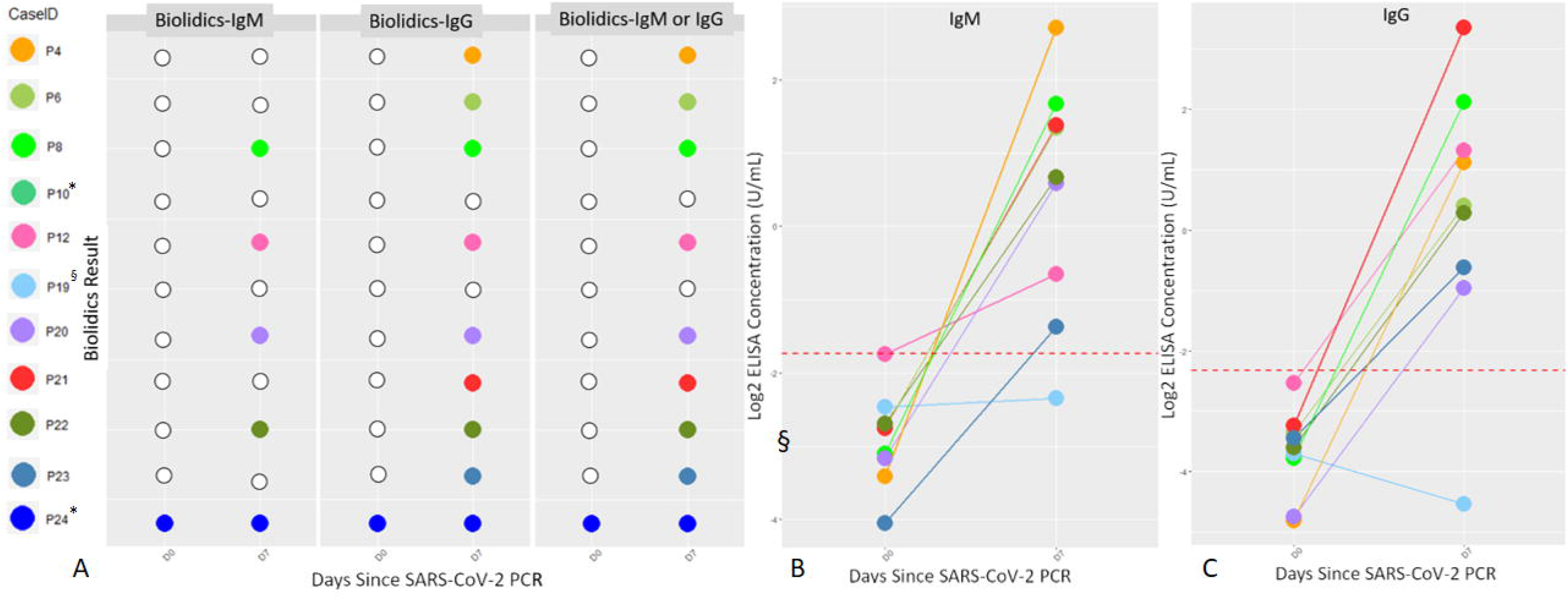
Sequential Antibody Detection. A. Samples of 11 patients with SARS-CoV-2 were tested by Biolidics at both D0 and D7 for the presence of IgM and IgG. Samples of 9 patients with SARS-CoV-2 were assessed by ELISA at D0 and D7 for the presence of IgM (B) and IgG (C). Dotted line represents ELISA concentration threshold of positivity (0.3 U/mL for IgM, 0.2 U/mL for IgG). * ELISA results not available for P10 and P24. §P19 was taking immunosuppressant therapy.

### Technical Agreement: Analytical Sensitivity and Specificity

Results of serologic LFI were correlated with results obtained by ELISA. ELISA was performed with plasma or serum samples from 15 patients with SARS-CoV-2 detected on nasopharyngeal swab and 24 negative control samples (Table 3A, 3B). Samples from 9 patients with SARS-CoV-2 were sequentially assessed at both D0 and D7, and samples of 6 patients with SARS-CoV-2 were assessed at D0 only.

**Table 3A.** Comparison of Biolidics and ELISA in detecting anti-SARS-CoV-2 IgM and IgG in patients with SARS-CoV-2

| Case ID | Biolidics - D0 |  | ELISA - D0 |  |  |  | Biolidics - D7 |  | ELISA - D7 |  |  |  |
| --- | --- | --- | --- | --- | --- | --- | --- | --- | --- | --- | --- | --- |
|  |  |  | IgM |  | IgG |  |  |  | IgM |  | IgG |  |
|  | IgM | IgG | Con.(U/mL) | Qualitative | Con.(U/mL) | Qualitative | IgM | IgG | Con.(U/mL) | Qualitative | Con.(U/mL) | Qualitative |
| P12 | ND | ND | 0.298273 | ND | 0.172371 | ND | D | D | 0.63587 | D | 2.500835 | D |
| P6 | ND | ND | 0.148562 | ND | 0.097747 | ND | ND | D | 2.531461 | D | 1.329731 | D |
| P8 | ND | ND | 0.116125 | ND | 0.073048 | ND | D | D | 3.188637 | D | 4.371472 | D |
| P4 | ND | ND | 0.093699 | ND | 0.035675 | ND | ND | D | 6.561597 | D | 2.169781 | D |
| P19* | ND | ND | 0.180726 | ND | 0.076687 | ND | ND | ND | 0.196576 | ND | 0.043024 | ND |
| P20 | ND | ND | 0.111488 | ND | 0.037183 | ND | D | D | 1.50561 | D | 0.515859 | D |
| P21 | ND | ND | 0.147533 | ND | 0.105664 | ND | ND | D | 2.60145 | D | 10.27846 | D |
| P22 | ND | ND | 0.154795 | ND | 0.082659 | ND | D | D | 1.586229 | D | 1.219473 | D |
| P23 | ND | ND | 0.060455 | ND | 0.09201 | ND | ND | D | 0.387017 | D | 0.653404 | D |
| P13 | D | D | 0.735776 | D | 0.279153 | D |  |  |  |  |  |  |
| P14 | D | ND | 0.484651 | D | 0.582676 | D |  |  |  |  |  |  |
| P15 | D | ND | 0.190855 | ND | 0.095461 | ND |  |  |  |  |  |  |
| P16 | D | D | 0.476458 | D | 0.382543 | D |  |  |  |  |  |  |
| P17 | D | D | 0.963219 | D | 0.569583 | D |  |  |  |  |  |  |
| P18 | D | D | 2.347554 | D | 2.011194 | D |  |  |  |  |  |  |

SARS-CoV-2
\* P19 was on immunosuppressant therapy after solid organ transplantation.

**Table 3B.** Comparison of Biolidics and ELISA in detecting IgM and IgG in patients without SARS-CoV-2

| Case ID | SARS-CoV-2 PCR | Biolidics |  | ELISA |  |  |  |
| --- | --- | --- | --- | --- | --- | --- | --- |
|  |  | IgM | IgG | IgM |  | IgG |  |
|  |  |  |  | Concentration, U/mL | Qualitative Result | Concentration, U/mL | Qualitative Result |
| N1 | ND | ND | ND | 0.265114 | ND | 0.056651 | ND |
| N2 | ND | ND | ND | 0.080361 | ND | 0.032590 | ND |
| N3 | ND | ND | ND | 0.072441 | ND | 0.047239 | ND |
| N4 | ND | ND | ND | 0.052037 | ND | 0.167129 | ND |
| N7 | Pre-COVID | D | ND | 0.175193 | ND | 0.141718 | ND |
| N8 | Pre-COVID | ND | ND | 0.074783 | ND | 0.051337 | ND |
| N9 | Pre-COVID | ND | ND | 0.105123 | ND | 0.061823 | ND |
| N10 | Pre-COVID | ND | ND | 0.096289 | ND | 0.066875 | ND |
| N11 | Pre-COVID | ND | ND | 0.108742 | ND | 0.051337 | ND |
| N12 | Pre-COVID | ND | ND | 0.203530 | ND | 0.104540 | ND |
| N13 | Pre-COVID | ND | ND | 0.195427 | ND | 0.031008 | ND |
| N14 | Pre-COVID | ND | ND | 0.117061 | ND | 0.175510 | ND |
| N15 | Pre-COVID | ND | ND | 0.287495 | ND | 0.111247 | ND |
| N16 | Pre-COVID | ND | ND | 0.067092 | ND | 0.105664 | ND |
| N17 | Pre-COVID | ND | ND | 0.112410 | ND | 0.163977 | ND |
| N18 | Pre-COVID | ND | ND | 0.172999 | ND | 0.106785 | ND |
| N19 | Pre-COVID | ND | ND | 0.096289 | ND | 0.153423 | ND |
| N20 | Pre-COVID | ND | ND | 0.101553 | ND | 0.048830 | ND |
| N21 | Pre-COVID | ND | ND | 0.243457 | ND | 0.112358 | ND |
| N22 | Pre-COVID | ND | ND | 0.136403 | ND | 0.096605 | ND |
| N23 | Pre-COVID | ND | ND | 0.463464 | D | 0.163977 | ND |
| N24 | Pre-COVID | D | ND | 0.307833 | D | 0.165028 | ND |
| N25 | Pre-COVID | ND | ND | 0.073218 | ND | 0.057957 | ND |
| N26 | Pre-COVID | ND | ND | 0.080361 | ND | 0.106785 | ND |

Of the 6 patients with SARS-CoV-2 assessed at D0 only, all 6 were positive for IgM or IgG antibodies by LFI and 5 were positive for IgM or IgG antibodies by ELISA (Table 3A). Samples of the 9 remaining PCR-positive patients were assessed at both D0 and D7. At D0, none of these samples were positive for IgM or IgG antibodies by LFI or ELISA, showing perfect concordance (Table 3A). At D7, 8 of 9 samples (89%) were positive for IgM and/or IgG antibodies by both LFI and ELISA. Four samples were negative for IgM by LFI on D7 and positive by ELISA. IgG at D7 showed perfect concordance between LFI and ELISA (Table 3A). The overall sensitivity of ELISA for detecting IgM, IgG, and IgM or IgG antibodies was 33% for all at D0 and 89% for all at D7 (Table 3C). LFI and ELISA were compared at both D0 and D7 for detecting IgM, IgG, and IgM or IgG (Figure 2, Figure 3). There was no significant difference between LFI and ELISA in any comparison except borderline significance for detecting IgM at D7 (p = 0.04; Table 3E). Correlation of MGH ELISA and Biolidics LFI was analyzed to establish a 95% confidence threshold indicating that at least 95% of positive results by Biolidics LFI correspond to antibody concentration of at least the confidence threshold on ELISA (Figure 3, black dotted line). Our data suggest that Biolidics LFI positivity can be used as a substitute for high level antibody levels by ELISA.

**Table 3C:**
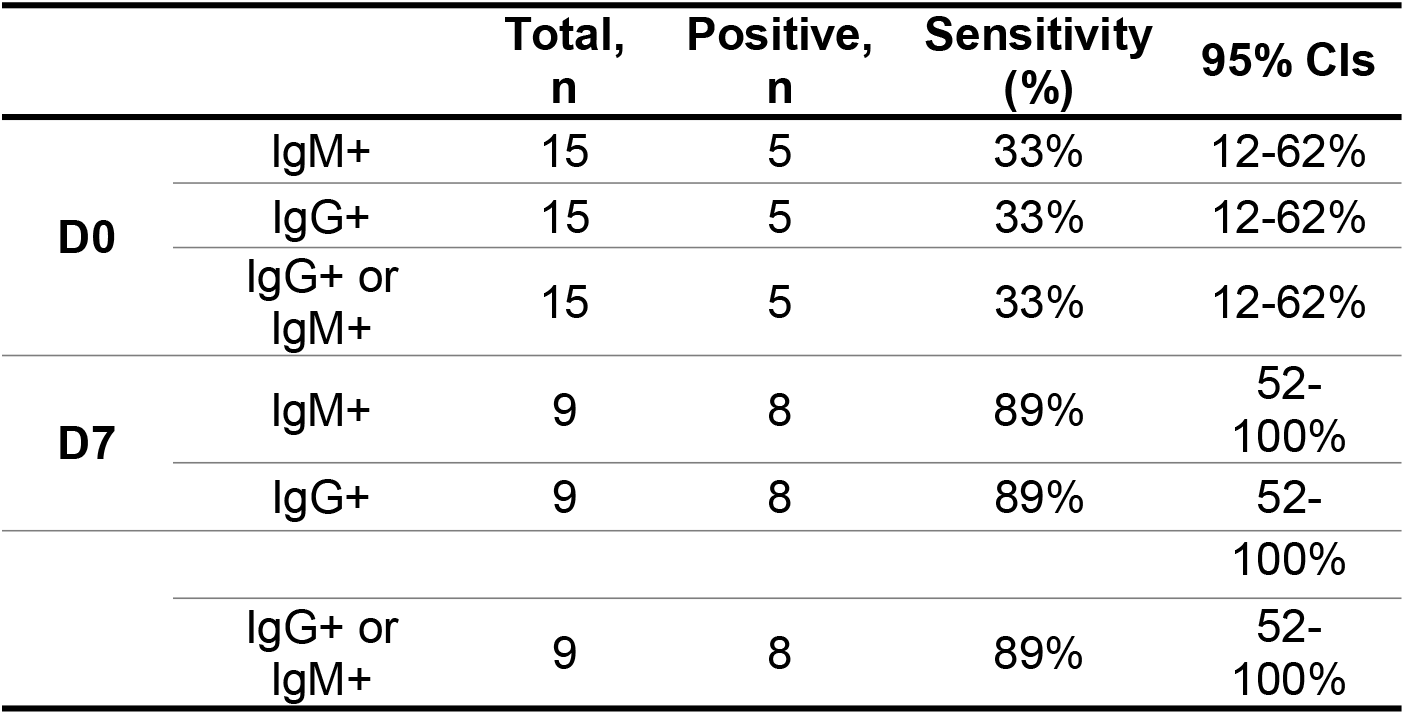
Percentage positivity for anti-SARS-CoV-2 IgM and IgG by ELISA in patients with SARS-CoV-2

**Table 3D:** Comparison of Biolidics and ELISA in detecting antibodies at D0 and D7

| PCR Date | Paired t-test | Biolidics Sensitivity | ELISA Sensitivity | p-value |
| --- | --- | --- | --- | --- |
| **D0** | IgM | 29% | 33% | 1.0 |
|  | IgG | 21% | 33% | 1.0 |
|  | IgM or IgG | 29% | 33% | 1.0 |
| **D7** | D7 IgM | 54% | 89% | 0.04 |
|  | D7 IgG | 88% | 89% | 1.0 |
|  | IgM or IgG | 92% | 89% | 1.0 |

\*D, detected; ND, not detected; D0, day 0 after PCR diagnosis; D7, day 7 after PCR diagnosis.

Of 24 negative plasma samples tested by ELISA, 22 (92%) were negative for both IgM and IgG antibodies by LFI, and 22 (92%) cases were negative for both IgM and IgG by ELISA. Two cases demonstrated IgM positivity by LFI. Interestingly, 2 cases also demonstrated positivity for IgM by ELISA. One case showed IgM by both ELISA and LFI, while 1 case each demonstrated IgM by LFI or by ELISA (Table 3B).

**Figure 3.**
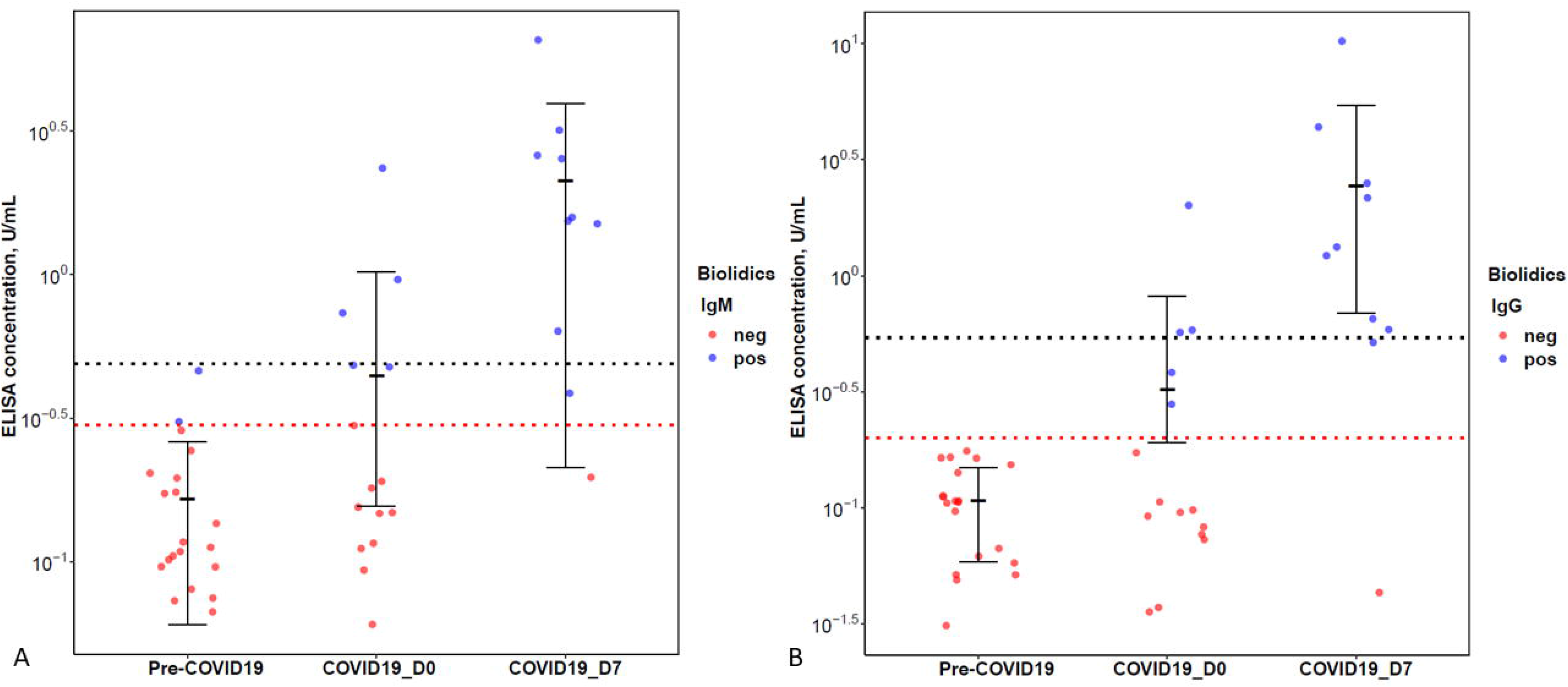
A. Comparison of Biolidics results and ELISA concentration in samples of patients from the pre-COVID19 era, patients with COVID19 tested at D0, and patients with COVID19 tested at D7. (A) IgM and (B) IgG. Red dotted line represents corresponding cut-off values based on ELISA standard curve, 0.3 U/mL for IgM and 0.2 U/mL for IgG. Black dotted line represents the 95% confidence threshold, such that 95% of results that are positive result by Biolidics have an antibody concentration by ELISA higher than or equal to the established threshold.

### Matrix Validation: Performance of Biolidics LFI using Venous Plasma and Whole Blood

Results of serologic LFI were compared using both D7 plasma and D7 venous whole blood samples from 22 patients with SARS-CoV-2 detected on nasopharyngeal samples (Table 4A; Figure 4A, 4B). With D7 plasma samples, 21 of 22 patients (95%) tested positive for antibodies (IgM and/or IgG). The same 21 patients tested positive for antibodies using D7 venous whole blood, indicating no significant difference between sample types for detecting IgM and IgG (p = 0.33, p = 1.00; Table 4B). Plasma and whole blood results for IgM and IgG separately were equivalent with one exception. In one patient (P35), IgM and IgG were both positive with D7 plasma, while IgM was negative and IgG was positive with D7 venous whole blood (Table 4A).

**Figure 4.**
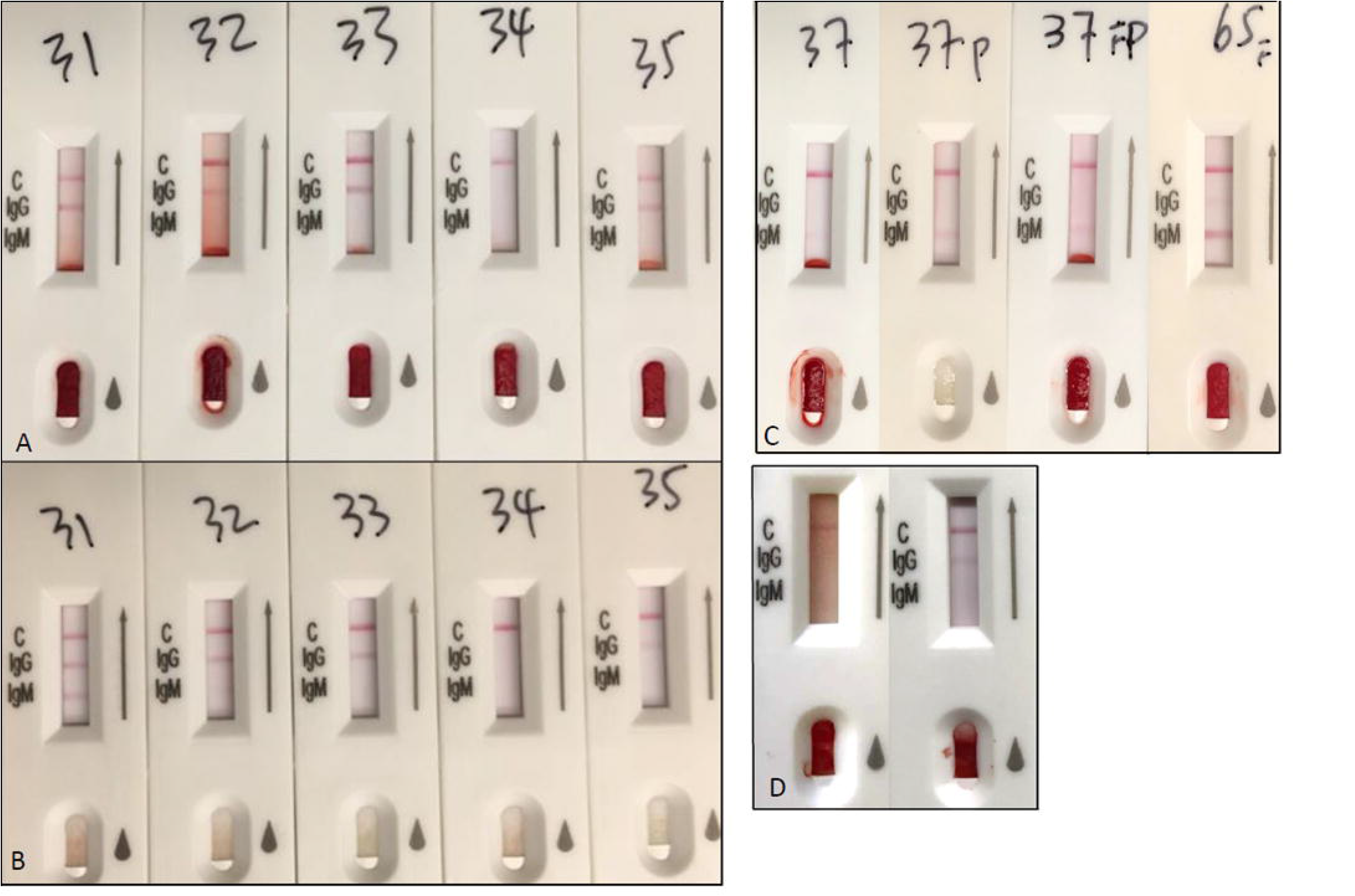
A-B. Biolidics LFI results in six paired venous plasma (A, top row) and whole blood (B, bottom row) samples showing concordant results. Test interpretation by numbered test kits: 31, IgM (+), IgG (+); 32, IgM (+), IgG (+); 33, IgM (-), IgG (+); 34, IgM (-), IgG (-); 35, IgM (-), IgG (+). C. Biolidics LFI results of (left to right) venous whole blood, venous plasma, capillary finger stick at 10 days after PCR diagnosis, and capillary finger stick at 16 days after PCR diagnosis. All 3 sample types at day 10 show IgM (+) and IgG (-), while capillary finger stick at day 16 shows IgM (+) and IgG (+). D. Biolidics LFI with capillary samples of two household members with simultaneous COVID-19. Left image shows IgM (-) and IgG(-) while right image shows IgM (-) and IgG (+).

**Table 4A:**
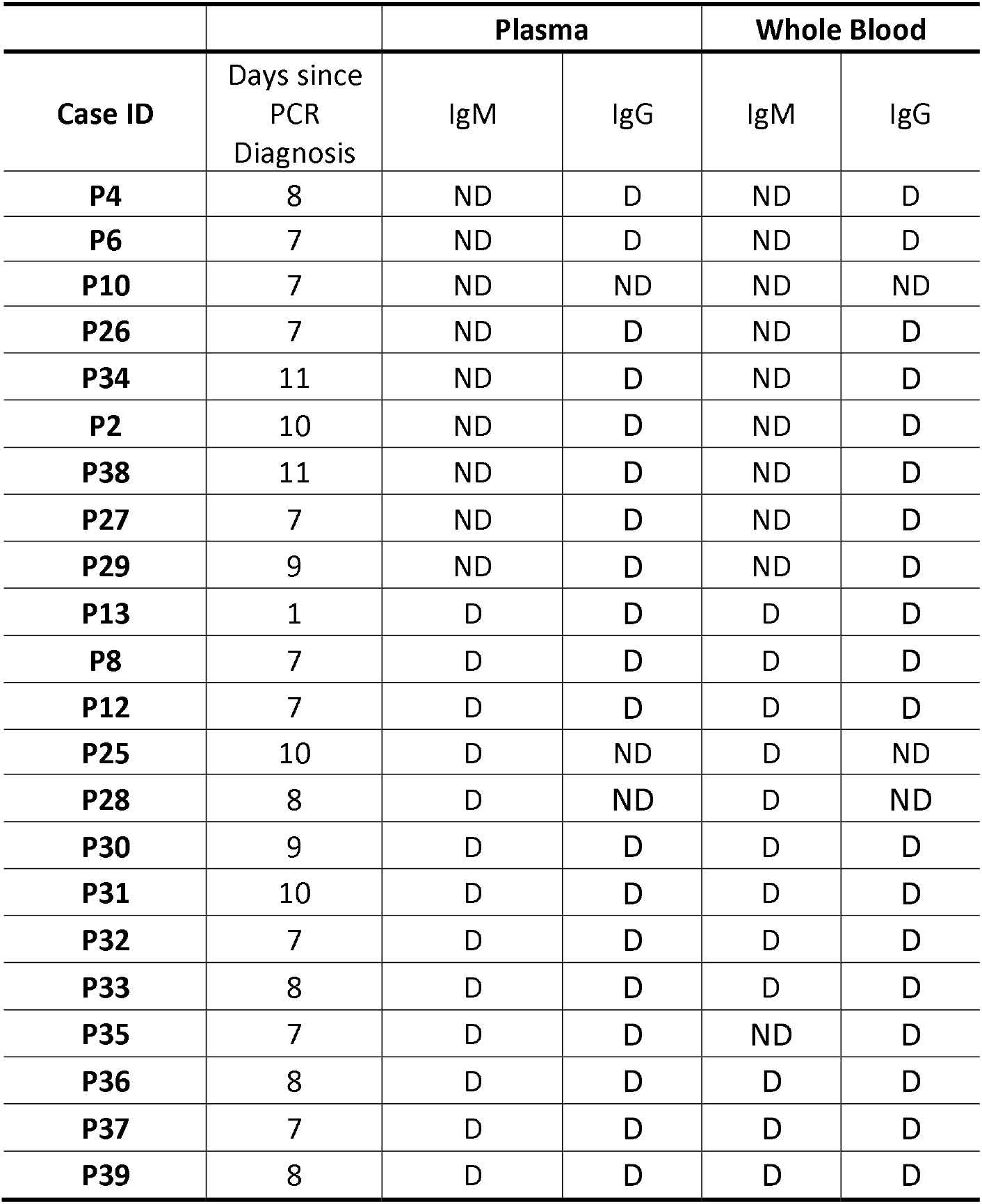
Performance of Biolidics in plasma and whole blood samples of patients with SARS-CoV-2

| Case ID | Days since PCR Diagnosis | Plasma |  | Whole Blood |  |
| --- | --- | --- | --- | --- | --- |
|  |  | IgM | IgG | IgM | IgG |
| P4 | 8 | ND | D | ND | D |
| P6 | 7 | ND | D | ND | D |
| P10 | 7 | ND | ND | ND | ND |
| P26 | 7 | ND | D | ND | D |
| P34 | 11 | ND | D | ND | D |
| P2 | 10 | ND | D | ND | D |
| P38 | 11 | ND | D | ND | D |
| P27 | 7 | ND | D | ND | D |
| P29 | 9 | ND | D | ND | D |
| P13 | 1 | D | D | D | D |
| P8 | 7 | D | D | D | D |
| P12 | 7 | D | D | D | D |
| P25 | 10 | D | ND | D | ND |
| P28 | 8 | D | ND | D | ND |
| P30 | 9 | D | D | D | D |
| P31 | 10 | D | D | D | D |
| P32 | 7 | D | D | D | D |
| P33 | 8 | D | D | D | D |
| P35 | 7 | D | D | ND | D |
| P36 | 8 | D | D | D | D |
| P37 | 7 | D | D | D | D |
| P39 | 8 | D | D | D | D |

**Table 4B:** Comparison of Plasma and Whole Blood Samples with Biolidics (Paired t-test)

| p-value |  |
| --- | --- |
| IgM | 0.33 |
| IgG | 1.00 |

### Performance of Biolidics LFI using Capillary Samples in Patients with SARS-CoV-2

In 14 patients who recovered from SARS-CoV-2, results of LFI with capillary (finger stick) samples were assessed at least two weeks after PCR confirmation (range, 18-46 days; median, 30.5 days) and symptom onset (range, 19-61 days; median, 32 days). Six patients tested positive for IgM, 12 tested positive for IgG, and 13 tested positive for IgM or IgG, for corresponding sensitivities of 43%, 86%, and 93%, respectively (Table 4C, 4D; Figure 4B). One patient was excluded from the sensitivity analysis due to concurrent diagnosis of an autoimmune disorder (systemic lupus erythematosus). Interestingly, this patient did not exhibit IgM or IgG antibodies on LFI.

**Table 4C:** Performance of Biolidics with capillary samples in patients with SARS-CoV-2

| Case ID | Symptom onset, days | Days since PCR diagnosis | Capillary |  |  |
| --- | --- | --- | --- | --- | --- |
|  |  |  | IgM | IgG | IgM or IgG |
| P40 | 19 | 18 | ND | D | D |
| P41 | 31 | 29 | ND | D | D |
| P42 | 31 | 30 | D | D | D |
| P43 | 28 | 25 | D | D | D |
| P44 | 32 | 28 | ND | D | D |
| P45 | 32 | 28 | ND | D | D |
| P46 | N/A* | 38 | ND | D | D |
| P47 | 36 | 35 | ND | D | D |
| P48 | 48 | 46 | ND | ND | ND |
| P49 | 48 | 45 | ND | D | D |
| P50 | 27 | 26 | D | D | D |
| P51 | 61 | 31 | D | D | D |
| P52 | 33 | 31 | D | D | D |
| P54 | 37 | 36 | D | ND | D |

\* Asymptomatic patient

**Table 4D:** Sensitivity of IgM and IgG detection with capillary sample in patients with SARS-CoV-2

| Symptom Onset | IgM+ | IgG+ | IgG+ or IgM+ |
| --- | --- | --- | --- |
| >14 days | 43% | 86% | 93% |
|  | (6/14) | (12/14) | (13/14) |
| 95% CIs | 16-68% | 52-96% | 60-98% |

\*D, detected; ND, not detected.

### Discussion

The COVID-19 pandemic has rapidly altered the landscape of clinical testing validation and regulatory oversight. With limited FDA oversight of serologic testing and variability of assays, rigorous internal validation has become paramount. This study validated the use of the commercially available 2019-nCoV IgG/IgM Detection Kit (Colloidal Gold) (Biolidics Ltd.) for detecting anti-SARS-CoV-2 IgM and IgG antibodies using plasma and whole blood samples from patients with PCR-confirmed SARS-CoV-2. Patient results were compared to those of negative control samples from hospitalized patients with negative SARS-CoV-2 PCR results as well as control samples archived before the emergence of SARS-CoV-2. We also sought to establish the sensitivity of the Biolidics test kit using capillary blood obtained via a finger stick, as LFI tests have potential utility in community testing, point of care testing, or even self-testing. Overall, we show that the Biolidics LFI kit has clinical sensitivity of 92% at 7 days after PCR diagnosis of SARS-CoV-2. Test specificity was 92% for IgM and 100% for IgG. Results of the LFI are similar to those obtained with gold standard ELISA testing, providing evidence that robust LFI tests can have utility comparable to that of ELISA with additional benefits of easy testing and low cost. There was no significant difference in detecting IgM and IgG with Biolidics LFI and ELISA at D0 and D7 (p=1.00), except for marginally significant detection of IgM at D7 (p=0.04). Nevertheless, IgM detection is more prone to false positive results by both ELISA and LFI.

Results obtained from plasma and whole blood samples were compared and show reproducibility between sample types, which removes the need for plasma or serum preparation. Furthermore, we show that capillary whole blood obtained by finger stick shows comparable sensitivity for detecting anti-SARS-CoV-2 IgM and IgG antibodies as venous blood samples. This provides an opportunity for community-based testing, rapid point of care testing, and potentially self-testing for the presence antibodies, similar to glucose self-monitoring. While ELISA remains a gold standard for establishing the level of antibodies, we show that strong correlation between LFI and ELISA can provide an estimate of antibody levels between closely correlated clinical assays. This close correlation decreases the need for additional testing by ELISA in settings where the financial cost of ELISA is a limiting factor prohibitive to broad serologic screening.

Serology provides important complementary information to PCR testing that is useful to evaluate the immunity status of a patient. Production of specific IgM antibodies is one of the body’s first lines of defense against viruses; IgG antibodies are produced 1-2 weeks later and provide long-term immunity. The presence of IgM antibodies can be used to indicate recent exposure, while IgG antibodies indicate previous exposure and often portend immunity [6]. In our study, detection of IgM was less specific than IgG detection by both Biolidics LFI and ELISA, and sensitivity for detection of IgM and IgG antibodies increased substantially with time. The most sensitive LFI results were obtained when both IgM and IgG were considered more than 7 days after PCR testing. Both LFI and ELISA at D7 showed positive IgM and/or IgG results for 8 of 9 sequentially tested patients. One patient did not show IgM or IgG antibodies on LFI or ELISA at D7; this patient previously underwent a solid organ transplant and was taking immunosuppressant therapy (Figure 2). Taken together, our results indicate that IgM and IgG serologic testing must be evaluated within the proper clinical context, and serial serologic assessment or follow-up testing with a more sensitive method assay may be useful in some circumstances.

While RT-PCR remains the gold standard for acute diagnosis of SARS-CoV-2, serologic tests provide supplementary diagnostic information and are more practical for use in large-scale screening [7-9]. Molecular-based PCR testing requires skilled technicians, relatively invasive sample collection, expensive technology, high complexity laboratories, and supplies that were in short supply due to high global demand. These molecular tests are useful for symptomatic patients who require triage and treatment, but antibody detection may be more useful for population-wide screening protocols. ELISA, the gold standard for antibody detection and quantification, requires trained technicians and relatively complex laboratory procedures that are difficult to quickly scale. In contrast, rapid LFI antibody tests are easy to use, require minimal training for performance and interpretation, are scalable for use in population-wide screening protocols, and are not limited to a laboratory setting.

In our study, use of the 2019-nCoV IgG/IgM Detection Kit (Colloidal Gold) did have important limitations. First, detection of antibodies increased from 29% at D0 to 92% at D7, indicating that sensitivity increased significantly with time. This is imperative to consider when implementing large-scale screening protocols. Repeat testing may be necessary to determine patients who have developed antibodies if they were first tested soon after exposure to the virus. This is especially true for asymptomatic carriers who never show symptoms but may still develop antibodies. Further large-scale longitudinal studies are needed to determine the expected timeline of antibody development in SARS-CoV-2. It is also imperative to determine whether long-term antibody production is maintained and whether antibodies provide effective immunity against SARS-CoV-2. Interestingly, we identified two patients in our capillary validation cohort who had similar symptoms of PCR-confirmed SARS-CoV-2 at the same time who lived in the same household. One patient had IgG detected by LFI over one month from diagnosis, while the other household member was negative for both IgM and IgG antibodies (Figure 4C), indicating possible interpersonal variability in the kinetics of developing SARS-CoV-2 antibodies. Further studies are warranted to evaluate inter-individual variability in the timing and strength of antibody response to the same strain of SARS-CoV-2. Finally, the 2019-nCoV IgG/IgM Detection Kit (Colloidal Gold) identified IgM antibodies in 2 of 20 negative controls archived before the emergence of SARS-CoV-2. However, ELISA cut-off values obtained from standard curves also detected IgM antibodies in 2 negative controls, resulting in the same specificity for IgM across Biolidics LFI and ELISA in this study. Importantly, neither LFI nor ELISA identified IgG antibodies in negative control specimens, suggesting that IgG is more accurate in identifying prior SARS-CoV-2 exposure. IgM antibody results should be interpreted in the proper clinical context, and repeated serology testing or immediate PCR testing to exclude asymptomatic infection may be warranted with detection of IgM alone.

In summary, rapid serology allows an effective tracking method for asymptomatic carriers and patients with mild disease who do not require sensitive molecular-based diagnosis to guide acute care. Here, we validated the use of Biolidics 2019-nCoV IgG/IgM Detection Kit (Colloidal Gold) and showed consistent results across multiple sample types compared to ELISA. However, the large variability of LFI tests requires that each test is either validated in-house or that proper FDA review and approval is conducted. From an interpretational standpoint, internal validation is recommended as different LFI tests may display different intensity of positive bands. Recording images and utilization of image software or artificial intelligence based interpretation of the results may also decrease inter-observer variability, which is particularly important in the setting of population-wide self-testing. Large-scale population based antibody screening with validated testing platforms will provide additional understanding of the human to human transmission rate, prevalence, incidence, and mortality of SARS-CoV-2, which remain largely uncertain in the rapidly evolving global landscape. Independently validated serology tests will be integral to obtaining this information.

## Data Availability

There is no publicly available data associated with this study.

## Acknowledgements

Y. Feng’s research is partially supported by NSF CAREER Grant DMS-2013789. The Authors would like to thank the NYU Center for Biospecimen Research and Development (CBRD) for help with sample collection and preparation, and healthcare providers and patients whose samples contributed to this study.

## References

1. Tu, Y.F., et al., A Review of SARS-CoV-2 and the Ongoing Clinical Trials. Int J Mol Sci, 2020. 21(7).

2. World Health Organization. WHO Characterizes COVID-19 as a Pandemic. 2020 [cited 2020 April 23]; Available from: https://www.who.int/emergencies/diseases/novel-coronavirus-2019/events-as-thev-happen.

3. Center for Systems Science and Engineering (CSSE) at Johns Hopkins University. COVID-10 Dashboard. 2020 23 April, 2020 at 8:31:22 AM [cited 2020 23 April]; Available from: https://coronavirus.ihu.edu/map.html.

4. Centers for Disease Control and Prevention. Cases of Coronavirus Disease (COVID-19) in the US. 2020 [cited 2020 23 April]; Available from: https://www.cdc.gov/coronavirus/2019-ncov/cases-updates/cases-in-us.html.

5. Cheng, M.P., et al., Diagnostic Testing for Severe Acute Respiratory Syndrome-Related Coronavirus-2: A Narrative Review. Ann Intern Med, 2020.

6. di Mauro, G., et al., SARS-Cov-2 infection: Response of human immune system and possible implications for the rapid test and treatment. Int Immunopharmacol, 2020. 84: p. 106519.

7. Babiker, A., et al., SARS-CoV-2 Testing. Am J Clin Pathol, 2020.

8. Tang, Y.W., et al., The Laboratory Diagnosis of COVID-19 Infection: Current Issues and Challenges. J Clin Microbiol, 2020.

9. Shen, M., et al., Recent advances and perspectives of nucleic acid detection for coronavirus. J Pharm Anal, 2020.

